# Trends of non-tuberculous mycobacterial disease in a low-tuberculosis prevalence setting

**DOI:** 10.1101/2023.09.24.23296023

**Authors:** Biplob Kumar Mohanty, Tomas Mikal Lind Eagan, Bernt Bøgvald Aarli, Dag Harald Skutlaberg, Tehmina Mustafa

**Author notes:** Correspondence Biplob Kumar Mohanty, University of Bergen, Department of Global Public Health and Primary Care Centre for International Health, 5020 BERGEN NORWAY.

## Abstract

**Background:** Limited data are available regarding factors associated with initiation of treatment and treatment outcomes after diagnosis of non-tuberculous mycobacteria (NTM) pulmonary infection and disease.

**Objective:** To investigate trends of NTM pulmonary infections, patient characteristics and factors associated with initiation of treatment and treatment outcomes in patients with NTM pulmonary infection and disease.

**Methods:** We evaluated 154 patients with NTM pulmonary infection, identified by having at least one record coded with ICD-10 A31.0 at Haukeland University Hospital in Bergen, Norway, from 2000 to 2021. A univariate and multivariate binary logistic regression was carried out to find the odds of factors associated with the initiation of treatment and treatment outcomes.

**Results:** 70% of the patients were older than 65 years. 49 % of patients had pulmonary comorbidity and the three most common symptoms were cough, dyspnoea, and weight loss. The most frequently observed mycobacterial species was *M. avium complex* (MAC), followed by *M. malmoense*, and *M. abscessus*. There was a decreasing trend in NTM pulmonary infection and NTM pulmonary disease from the year 2000 to 2014, while an increase was observedfrom 2015 to 2019. A total of 72 (47%) patients received antibiotic treatment. Patients with high symptom scores, those below the age of 65, and those with MAC infection had more than three times the odds of receiving antibiotic treatment (P = 0.006, P = 0.006, and P = .005 respectively). Of 72 patients who received treatment, 53 (74%) had a favourable response and culture conversion. 17 (32%) of them had a relapse. Out of 82 patients who did not receive treatment, 45 (55%) had spontaneous culture conversion. 8 (18%) of them had a relapse. No factor was identified to be significantly associated with a favourable treatment response including the time taken to start treatment or presence of pulmonary cavities.

**Conclusion:** Favourable response to treatment was seen in 74% patients whereas spontaneous culture conversion was seen in 55% of non-treated patients. Factors associated with favourable treatment response were not found.

## Introduction

Non-tuberculous mycobacterial (NTM) infections are a public health problem worldwide. Despite being overshadowed by infections from other members of the *Mycobacteriaceae* family, such as *Mycobacterium tuberculosis* and *Mycobacterium leprae*, NTM infections are associated with considerable mortality and morbidity [1, 2]. The incidence of NTM infections is particularly high in the elderly population, and it is projected to continue to rise in the coming years due to the increasing number of elderly individuals [3].

NTM infections can cause a range of diseases, including non-tuberculous mycobacterial pulmonary disease (NTM-PD). Some of the more common NTM species known to cause NTM-PD are *Mycobacterium avium complex* (MAC), *Mycobacterium kansasii, Mycobacterium xenopi, Mycobacterium abscessus,* and *Mycobacterium malmoense* [4]. These infections typically occur as comorbidities in patients with underlying respiratory diseases such as chronic obstructive pulmonary disease (COPD), bronchiectasis, and cystic fibrosis (CF) [5]. However, diagnosis of NTM infections can be challenging, and patients often face a lengthy time to diagnosis or misdiagnosis, leading to a poor long-term outcome. Information on the prevalence of NTM infections is usually lacking as NTM infections do not require official notification. However, there is a need to investigate the trends and characteristics of NTM infections in specific geographic areas to provide information for the timely diagnosis and treatment of these infections.

This study aimed to investigate the trends and characteristics of NTM pulmonary infections and NTM-PD over the past two decades at a tertiary care hospital in western Norway. We aimed to identify factors associated with initiating treatment for NTM infection and treatment outcome.

## Methodology

### Study setting and design

The study is a retrospective cohort investigation carried out at Haukeland University Hospital, Bergen, Norway. To identify eligible cases, all inpatient and outpatient records were searched for the International Classification of Diseases, 10th Revision (ICD-10) code A31.0 (pulmonary mycobacterial infection) between the years 2000 and 2021. Patients diagnosed with NTM pulmonary infection and NTM-PD that met the current American Thoracic Society (ATS) criteria were included in the study.

**Figure 1** presents the selection process for the study sample. Two hundred and twenty-one potential patients with NTM pulmonary infection were identified by having at least one record coded with A31.0. NTM pulmonary infection was defined as a positive culture for NTM from two separate expectorate sputum samples or positive culture from one bronchoalveolar lavage. NTM-PD was defined as NTM pulmonary infection with pulmonary or systemic infection, symptoms, and nodular or cavitary opacities on chest X-ray or chest computer tomography (CT) scan. Among the 100 deceased patients records with NTM pulmonary infection from 83 patients with confirmed NTM pulmonary infection were available. Thirty-five patients who were alive and under 70 years old did not provide consent and were excluded. For alive patients above the age of 70, 42 records were available. The final study sample included 154 patients.

**Figure 1.**
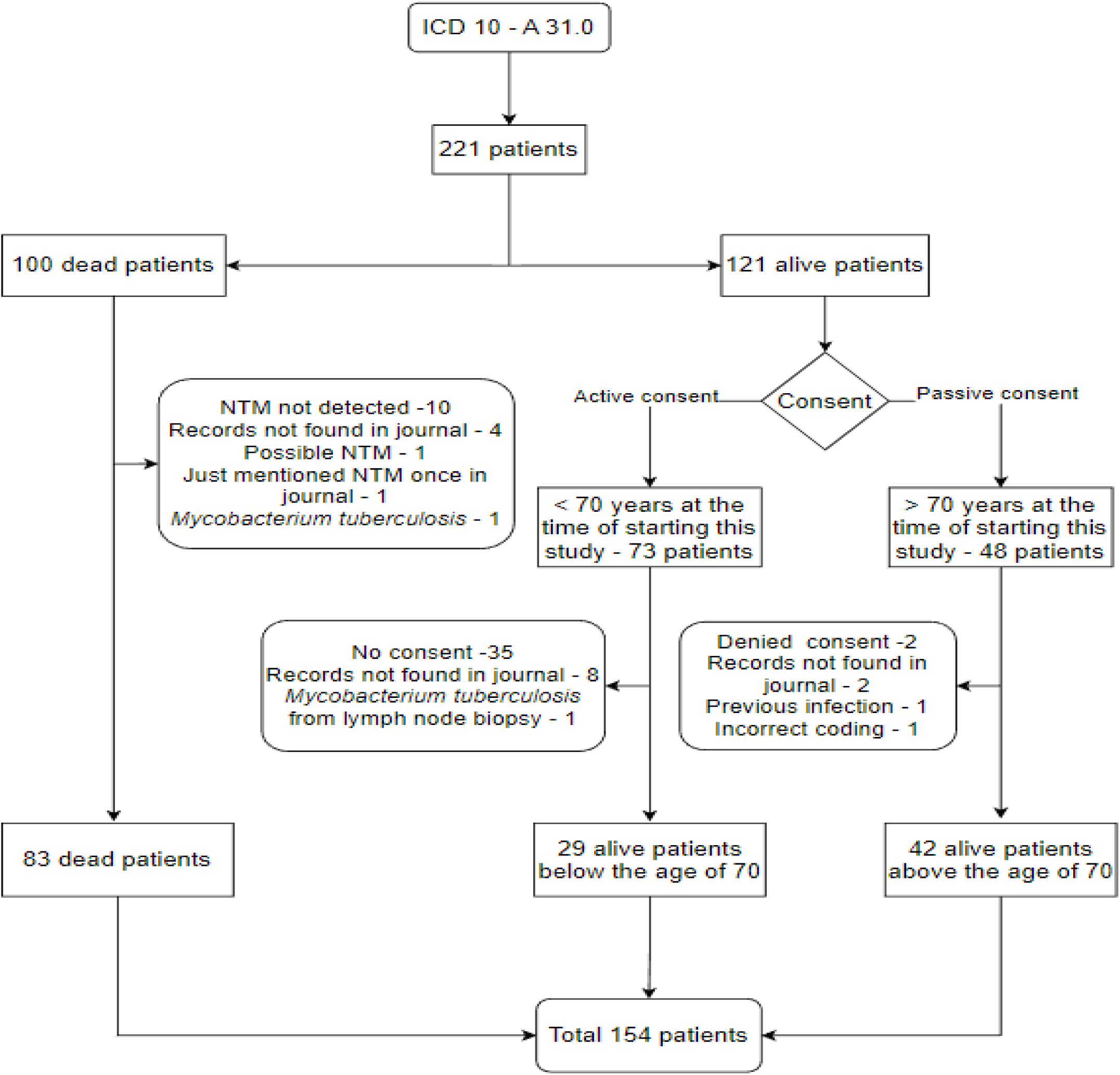
Flow chart showing inclusion and exclusion of patients in the study.

Relevant data were collected from the electronic medical records of the Department of Thoracic Medicine, Haukeland University Hospital. The variables collected included sex, age, presence of pulmonary comorbidities, symptoms, microbiological findings, radiological findings (both chest X-ray and chest CT scan), treatment history, and treatment outcome. Six symptoms of particular interest were recorded: fever, cough, haemoptysis, night sweats, dyspnoea, and weight loss. A symptom score was calculated, which was defined as a sum of the total number of symptoms where a score of 1 is given for each symptom. A score of 3 or more was considered a high symptom score. A favourable response to treatment or culture conversion was defined as at least two consecutive negative mycobacterial cultures from respiratory samples.

### Data management and analysis

The collected data were analysed using IBM SPSS statistics software version 27. Binary logistic regression was used for univariate and multivariate analyses to identify factors associated with starting of treatment and factors associated with treatment outcome. The unadjusted odds ratio (OR) and 95% confidence interval (CI) for each variable were first calculated using univariate regression. Multivariate regression was then performed. All variables were included in the final model. The adjusted OR (aOR) and 95% CI was calculated for each included variable. A p-value ≤0.05 was considered statistically significant.

### Ethical clearance

Ethical clearance was obtained from the Regional Committee for Medical Ethics Western Norway, reference number: 282165. Active consent was obtained from patients below the age of 70, while passive consent was taken from patients above the age of 70. In the case of deceased patients, consent was not required. All data were de-identified to protect patients’ privacy (i.e., social security numbers, names, or other directly identifiable characteristics were removed), in accordance with local regulations.

## Results

The demographic and clinical characteristics of patients with NTM pulmonary infection and NTM-PD are presented in Table 1. 70% of the study sample were patients older than 65 years. The most commonly reported symptoms were cough (82%), dyspnoea (43%), and weight loss (33%). Pulmonary cavities were observed in 25% of the patients. Among the patients with NTM pulmonary infection and NTM-PD, 30% were found to have COPD and 19% had bronchiectasis. A total of six main groups/species causing NTM infections were identified, including *M. avium complex* (MAC), *M. malmoense*, *M. abscessus*, *M. gordonae*, *M. fortuitum*, and *M.* xenopi.

**Table 1.**
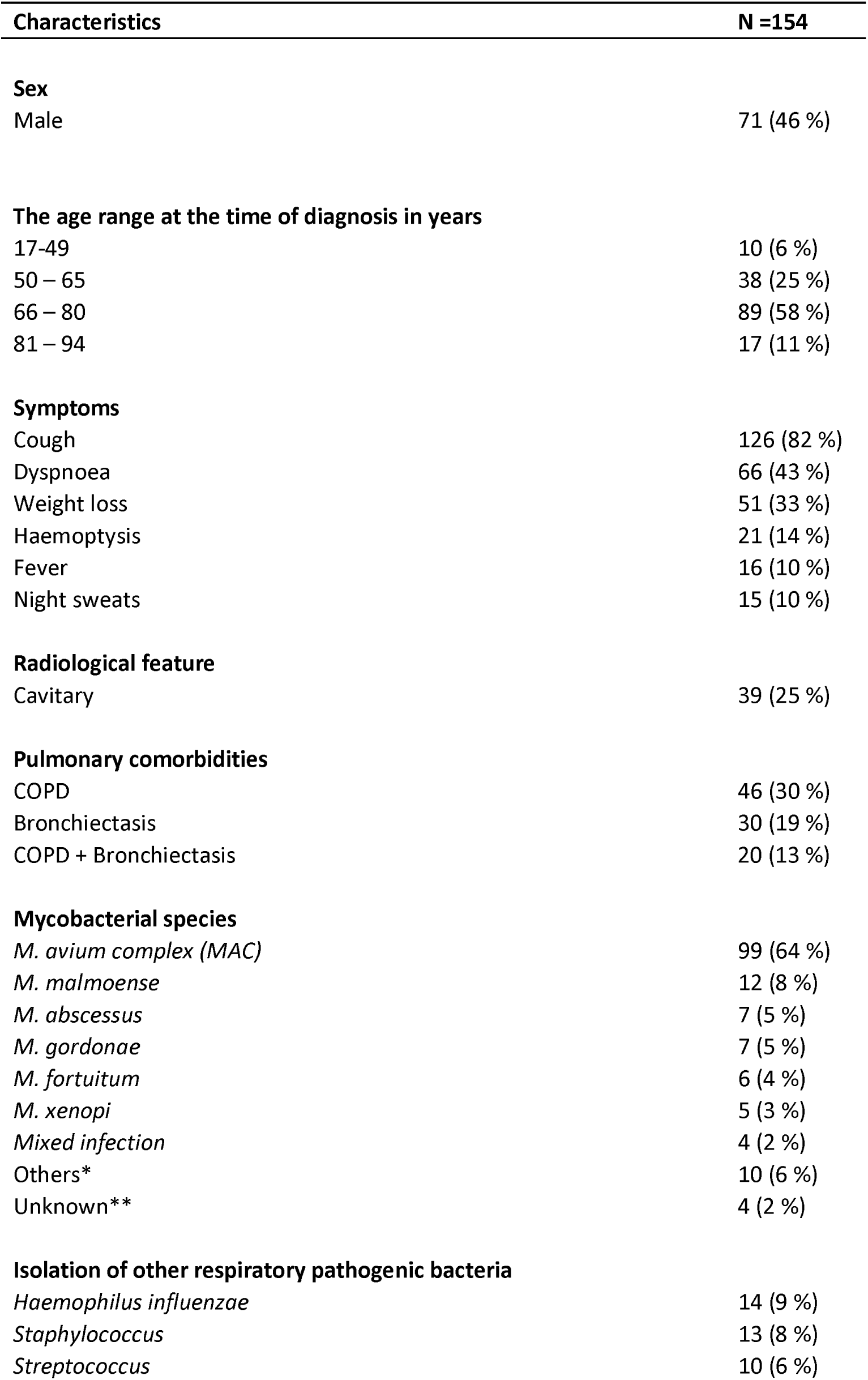
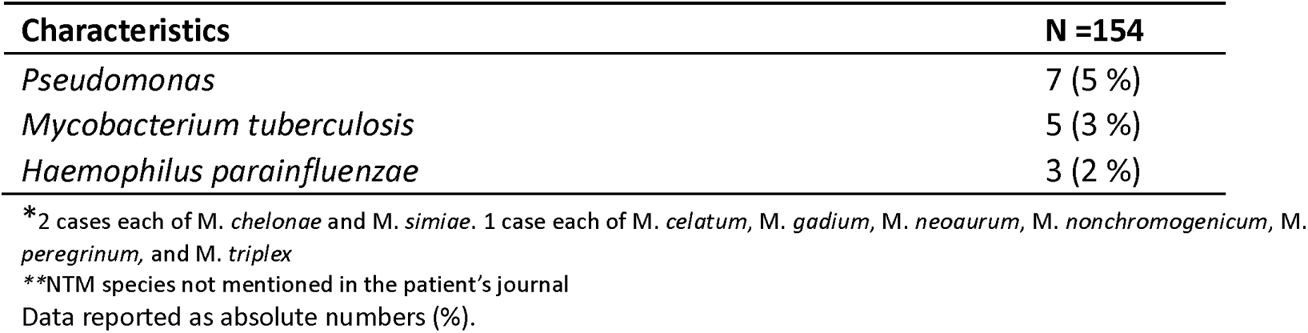
Demographic and clinical characteristics of patients with NTM pulmonary infection and NTM-PD.

The proportion of different NTM species in NTM pulmonary infections and NTM-PD over the past 22 years is presented in Figures 2.1 and 2.2 respectively. Figure 2.3 shows that the number of NTM infection cases declined every 5 years, with 48 (31%), 36 (23%), and 19 (12%) cases reported for the time periods 2000-2004, 2005-2009, and 2010-2014, respectively. There was an increase in the number of cases in the period 2015-2019, with 38 (25%) cases reported. The trend for NTM pulmonary disease followed a similar pattern. All species follow a similar trend across the years.

**Figure 2.1:**
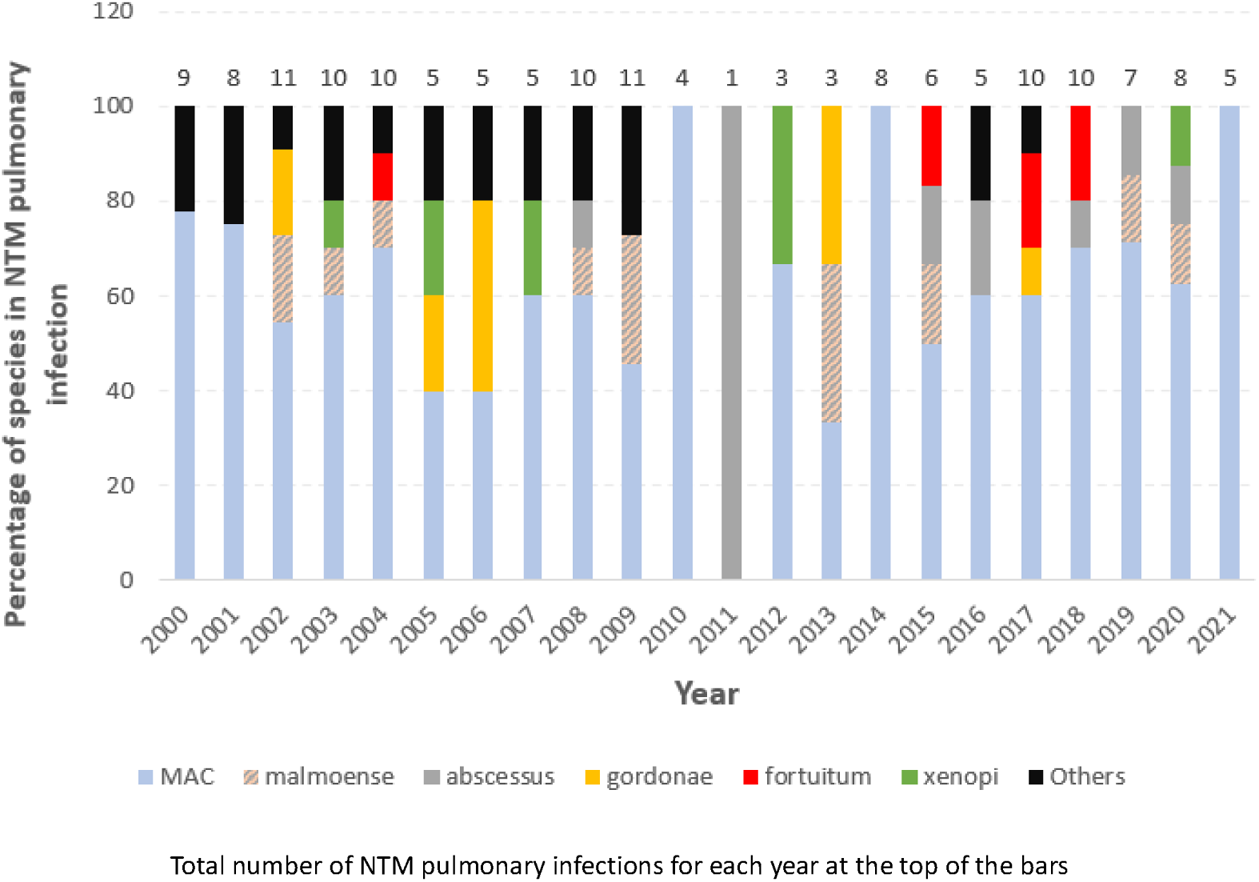
Proportion of different species in NTM pulmonary infection from the year 2000 to 2021.

**Figure 2.2:**
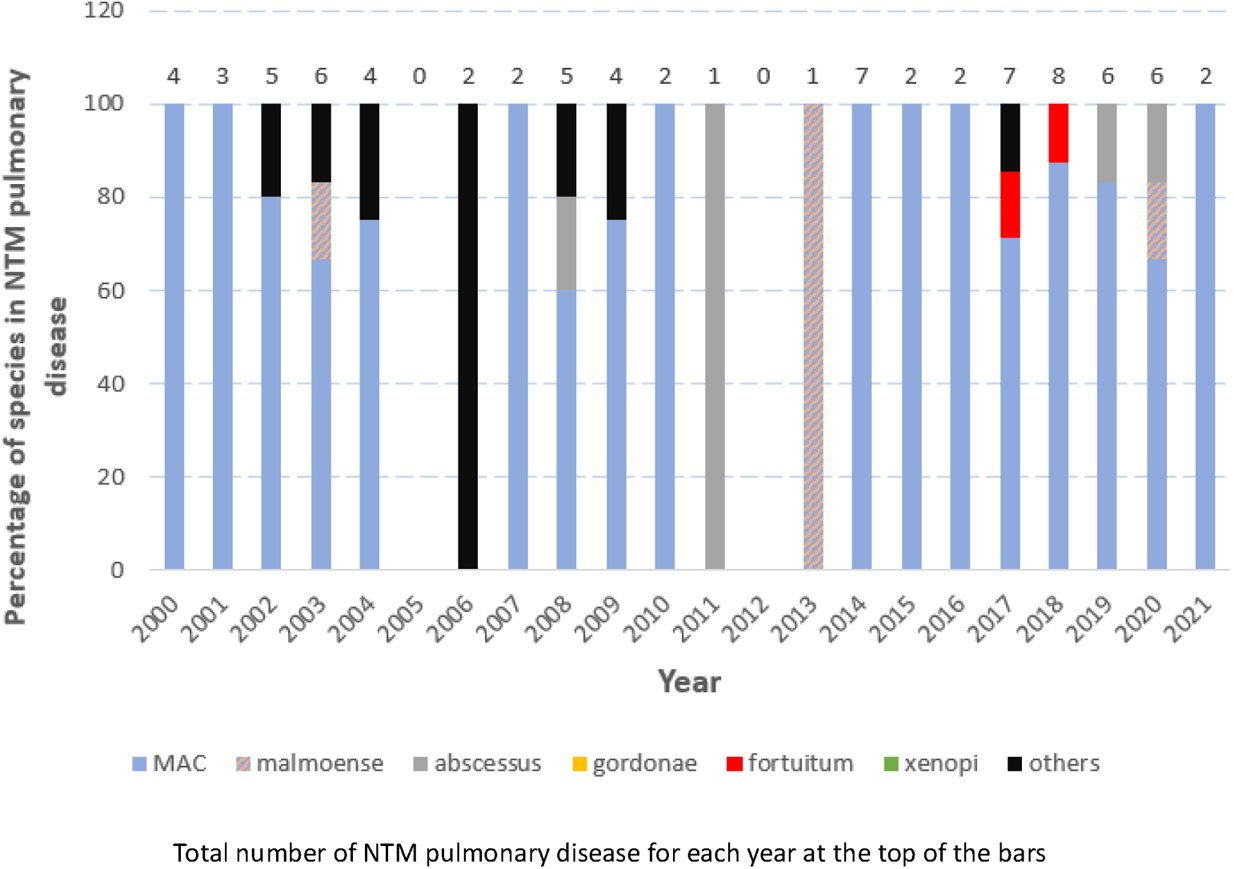
Proportion of different species in NTM pulmonary disease from the year 2000 to 2021.

**Figure 2.3:**
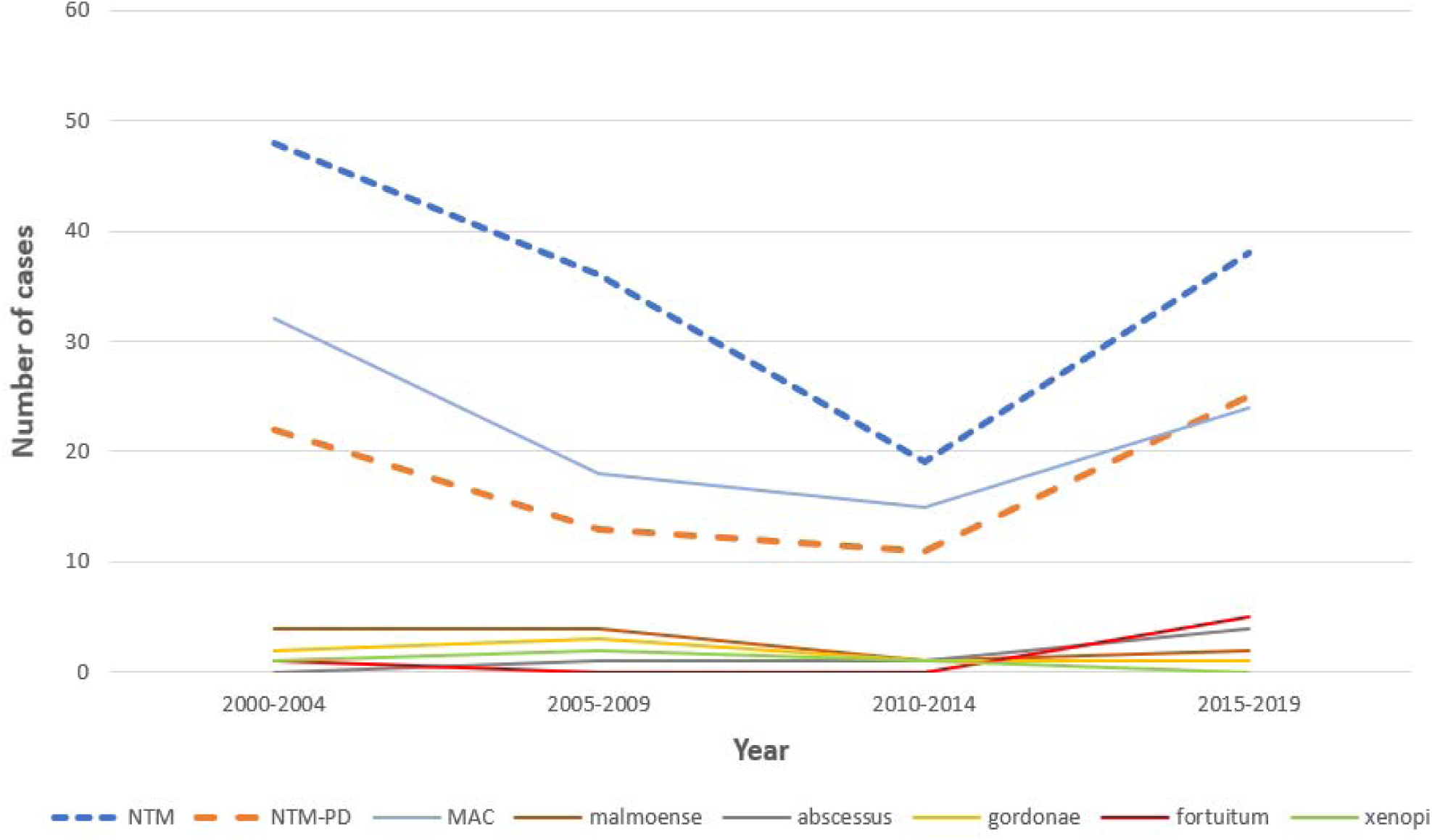
Trend of different species in NTM pulmonary infection in 5 years period from the year 2000 to 2019.

**Table 2** shows the clinical characteristics of patients with NTM pulmonary disease caused by different species. All species were associated with low symptom scores and predominantly non-cavitary disease. Apart from *M. gordonae*, infections with all species received treatment. All species showed a positive response to treatment in more than 60% of the cases except for *M. malmoense*, where a positive response to treatment was shown in only 25% of cases. In *M. malmoense* infections, relapse was seen in 100% of cases.

**Table 2:** Clinical characteristics of patients with NTM pulmonary infection caused by different species.

| Category | MAC<br>(N = 99) | M.<br>malmoense<br>(N = 12) | M.<br>abscessus<br>(N = 7) | M.<br>gordonae<br>(N = 7) | M.<br>fortuitum<br>(N = 6) | M.<br>xenopi<br>(N = 5) |
| --- | --- | --- | --- | --- | --- | --- |
| Age below 65 | 31 (31) | 5 (42) | 4 (57) | 1 (14) | 2 (33) | 2 (40) |
| Male | 43 (43) | 7 (58) | 1 (14) | 4 (57) | 3 (50) | 2 (40) |
| Cavities | 24 (24) | 5 (42) | 1 (14) | 1 (14) | 2 (33) | 2 (40) |
| High symptom score | 37 (37) | 4 (33) | 2 (29) | 0 (0) | 1 (17) | 2 (40) |
| Treatment given | 56 (57) | 4 (33) | 5 (71) | 0 (0) | 2 (33) | 2 (40) |
| Positive response to treatment* | 42 (75) | 1 (25) | 3 (60) | NA | 2 (100) | 2 (100) |
| Relapse** | 15 (36) | 1 (100) | 0 (0) | NA | 0 (0) | 1 (50) |

Data reported as n (%)
Percentage calculated by the formula $n \times 100/N$
\*N is equal to n of treatment given
\*\*N is equal to n of positive response to treatment

A total of 71% of patients with *M. abscessus* infection, 57% of patients with MAC infection, 40% of patients with *M. xenopi* infection, and 33% of patients with *M. malmoense* and *M. fortuitum* received treatment. Table 3 shows the factors associated with initiation of antibiotic treatment. Of the 154 patients with NTM pulmonary infection, 79 patients fulfilled criteria for NTM-PD. Treatment was started for both patients with infection and disease. In all, 72 (47%) patients received antibiotic treatment. Of these, 49 patients had NTM-PD and 23 patients had NTM pulmonary infection. Patients with high symptom scores, those below the age of 65, and those with MAC infection had more than three times the odds of receiving treatment (P = 0.006, P = 0.006, and P = .005 respectively). Thirty-one (56%) patients were started on treatment more than 6 months after diagnosis. The first guidelines on the treatment of NTM lung disease in 2007 did not impact the decision to start treatment.

**Table 3:**
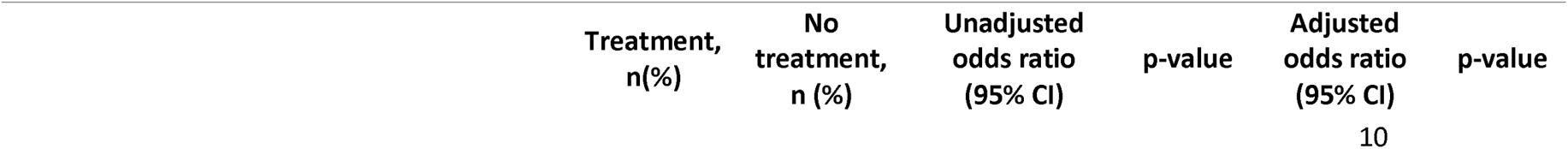
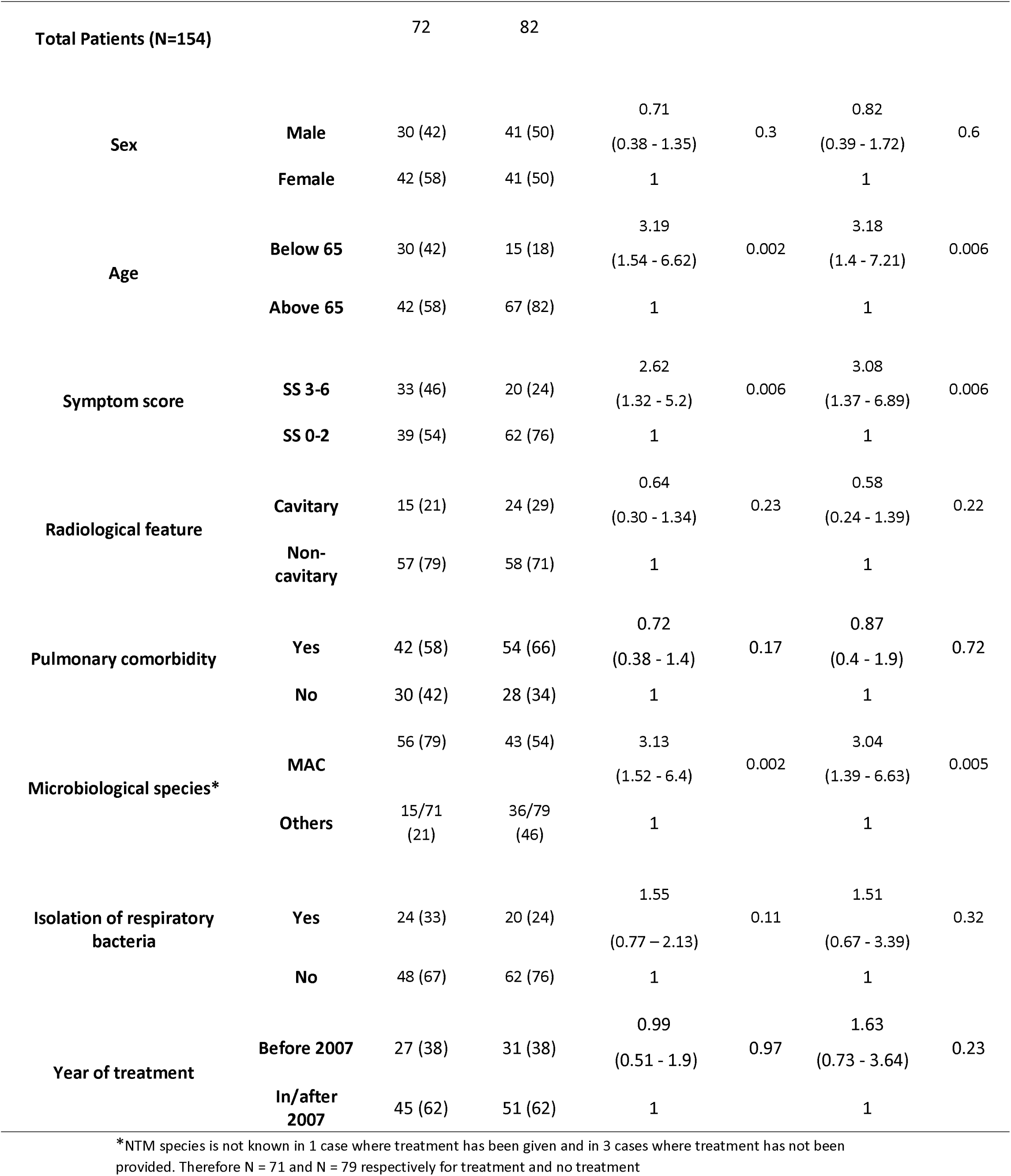
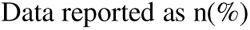
Factors associated with initiation of antibiotic treatment for eradication of non-tuberculous mycobacteria (NTM) among patients with NTM pulmonary infection.

Figure 3 shows the comparison of culture conversion between treatment and non-treatment groups. Out of 72 patients who received treatment, 53 (74%) had a favourable response and had culture conversion. 17 (32%) of them had a relapse. Out of 82 patients who did not receive treatment, 45 (55%) had spontaneous culture conversion. 8 (18%) of them had a relapse.

**Figure 3:**
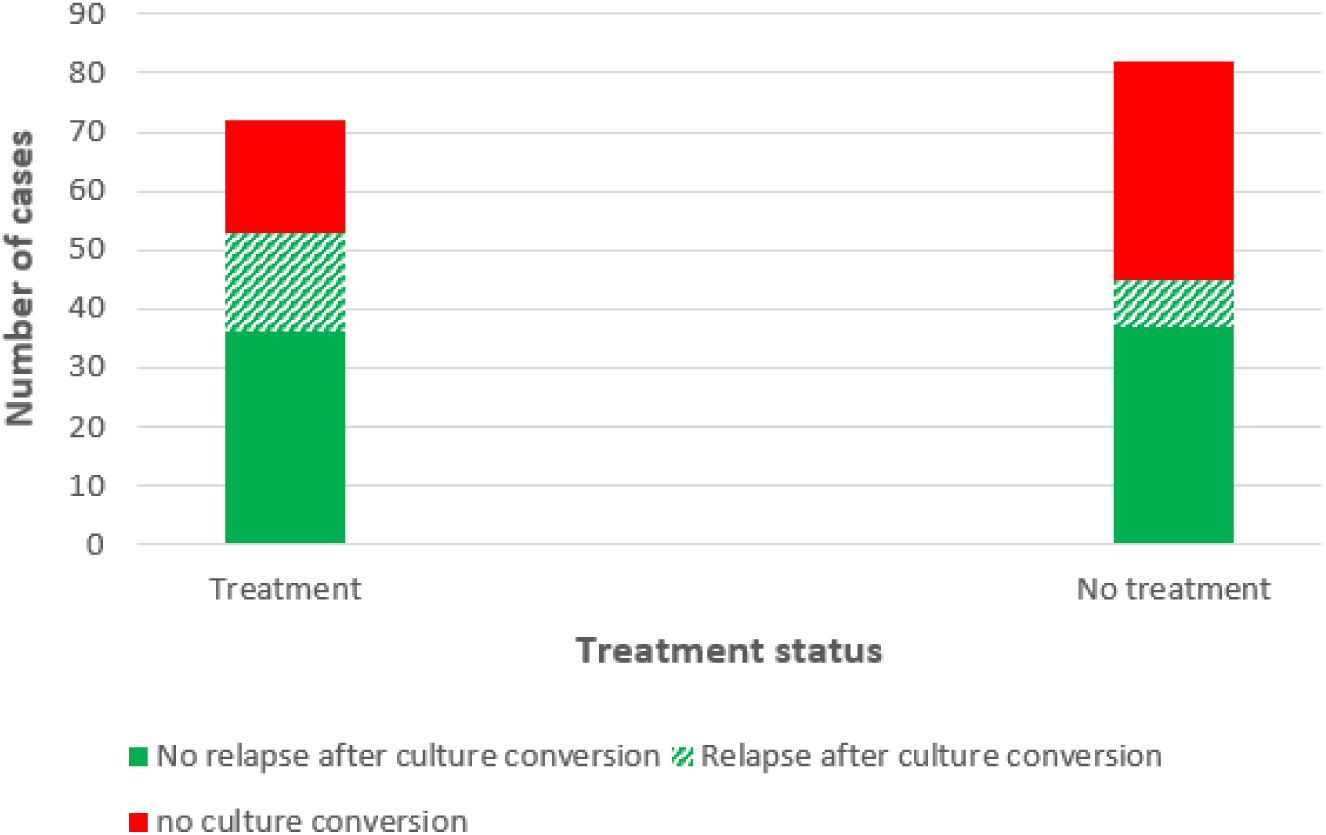
Comparison of culture conversion between patients with NTM pulmonary infection who received or did not receive treatment.

Table 4 shows the factors associated with favourable response to treatment. No factor was identified to be significantly associated with a favourable treatment response including the time taken to start treatment or presence of pulmonary cavities.

**Table 4:** Factors associated with favourable response to treatment among patients with NTM pulmonary infection.

|  |  | Favourable response to treatment, n(%) | No response to treatment, n (%) | Unadjusted odds ratio (95% CI) | p-value | Adjusted odds ratio (95% CI) | p-value |
| --- | --- | --- | --- | --- | --- | --- | --- |
| **Total patients (N)** |  | 53 (74) | 19 (26) |  |  |  |  |
| **Sex** | **Male** | 21 (40) | 9 (47) | 0.73<br>(0.25 - 2.1) | 0.56 | 0.85<br>(0.27 - 2.64) | 0.78 |
|  | **Female** | 32 (60) | 10 (53) | 1 |  | 1 |  |
| **Age** | **Below 65** | 20 (38) | 10 (53) | 0.55<br>(0.19 - 1.57) | 0.26 | 0.55<br>(0.17 - 1.72) | 0.3 |
|  | **Above 65** | 33 (62) | 9 (47) | 1 |  | 1 |  |
| **Symptom score** | **SS 3 – 6** | 25 (47) | 08 (42) | 1.22<br>(0.43 - 3.54) | 0.7 | 1.16<br>(0.36 - 3.72) | 0.8 |
|  | **SS 0 - 2** | 28 (53) | 11 (58) | 1 |  | 1 |  |
| **Radiological feature** | **Cavitary** | 10 (19) | 5 (26) | 0.65<br>(0.19 - 2.23) | 0.5 | 0.63<br>(0.16 - 2.39) | 0.49 |
|  | **Non-cavitary** | 43 (81) | 14 (74) | 1 |  | 1 |  |
| **Time taken for initiation of treatment\*** | **Less than 6 months** | 23 (55) | 8 (62) | 0.69<br>(0.34 - 1.41) | 0.31 | 0.73<br>(0.34 - 1.56) | 0.42 |
|  | **More than 6 months** | 19 (45) | 5 (38) | 1 |  | 1 |  |

|  |  | Favourable<br>response to<br>treatment,<br>n(%) | No response<br>to treatment,<br>n (%) | Unadjusted<br>odds ratio<br>(95% CI) | p-value | Adjusted<br>odds ratio<br>(95% CI) | p-value |
| --- | --- | --- | --- | --- | --- | --- | --- |
| Pulmonary<br>comorbidity | **Yes** | 28 (60) | 8 (53) | 2.08<br>(0.71 - 6.1) | 0.18 | 1.95<br>(0.58 - 6.50) | 0.28 |
|  | **No** | 25 (40) | 10 (47) | 1 |  | 1 |  |
| Hypertonic saline<br>inhalation | **Yes** | 19 (36) | 7 (37) | 0.96<br>(0.32 - 2.84) | 0.94 | 0.77<br>(0.23 - 2.54) | 0.67 |
|  | **No** | 34 (64) | 12 (63) | 1 |  | 1 |  |
| Microbiological<br>species** | **MAC** | 42 (81) | 14 (74) | 1.63<br>(0.5 - 5.29) | 0.42 | 1.69<br>(0.46 - 6.13) | 0.43 |
|  | **Others** | 10 (19) | 5 (26) | 1 |  | 1 |  |

\*Since information is not available on time to initiation of treatment in 17 patients, the value of N changes to N = 42 for favorable response to treatment and N = 13 for response to treatment.
\*\*In one patient with favorable outcome to treatment, species is not mentioned in journal of the patient. So, N = 52 for favorable response to treatment in this case.
Data reported as n(%)

Figure 3 shows the comparison of culture conversion between treatment and non-treatment groups. Out of 72 patients who received treatment, 53 (74%) had a favourable response and had culture conversion. 17 (32%) of them had a relapse. Out of 82 patients who did not receive treatment, 45 (55%) had spontaneous culture conversion. 8 (18%) of them had a relapse.

Table 4 shows the factors associated with favourable response to treatment. No factor was identified to be significantly associated with a favourable treatment response including the time taken to start treatment or presence of pulmonary cavities.

## Discussion

To our knowledge this is the first study to investigate the trends of NTM pulmonary infections, and factors associated with initiation of treatment and outcomes from a high-income and low TB-endemic setting of Western Norway over a period of 20 years. The major species observed were MAC, *M. malmoense*, and *M. abscessus*. The NTM infections showed a declining trend till 2014 following an increase in later years. This increasing trend could be attributed to better diagnostic modalities and increased focus on the detection of mycobacteria from the respiratory samples. Furthermore, increase in life expectancy with pulmonary comorbidities could also be a contributing factor, as indicated by an increased risk of getting NTM infection with increased age and presence of pulmonary comorbidity in the current study. Several studies have shown that age is a significant factor associated with the rise in incidence rates of NTM pulmonary disease among older populations, with higher increase in the annual prevalence in individuals aged ≥ 60 years than in those aged < 60 years [6,7,8]. Al-Houqani et al. [9] found that age is a significant contributor to the increase in incidence rates of NTM-PD, however accounting for less than a quarter of the total increase. This suggests that other factors may also play a role in the rise of NTM pulmonary disease incidence rates. One such factor is chronic lung diseases, including COPD, bronchiectasis, and cystic fibrosis, which are known predisposing factors for NTM pulmonary disease development [10]. In the present study, COPD was the most associated chronic lung disease (30%), followed by bronchiectasis (19%). Both diseases have increased in prevalence over the last 30 years, and are themselves associated with higher age. COPD and bronchiectasis both disrupt the normal architecture of the airways, resulting in chronic bronchitis, with disruption of the epithelial membranes and increases in mucus production [11]. This unfortunately is an ideal environment for bacterial colonization and overgrowth. In addition, several patents with COPD may have reduced general immunity due to age or poor nutrition. Patients with COPD are prone to develop cachexia [12]. Finally, many COPD patients use inhaled corticosteroids (ICS) to prevent COPD exacerbations, which are often of viral initiation. In some COPD patients, ICS treatment may also increase their risk of bacterial infections, including NTM pulmonary disease [13]. In cystic fibrosis, due to impaired mucociliary clearance there are viscous airway secretions which predisposes the patient to bacterial colonization and infection [14]. A systematic review and meta-analysis review of 95 studies have reported prevalence of NTM infection in cystic fibrosis to be 7.9% and increasing over time [15].

In our data, 47% patients received antibiotic treatment for eradication of NTM. Patients with high symptom scores had greater odds of receiving treatment, and the odds of receiving treatment were much higher when the patient was below the age of 65. Interestingly, the presence of a cavity did not have a major impact on the initiation of treatment. In a study, it was found that patients had increased odds of receiving treatment in case of cavitation on CT imaging, presence of night sweats, and weight loss [16]. One study from South Korea demonstrated that long-term treatment success rate decreased with age, particularly in patients aged=≥80 years. Whereas the rate of adverse drug reactions requiring discontinuation of treatment increased with age, and the number was twice as high in patients aged=≥80 years than in those aged=<50 years [17]. These findings support the decision to treat relatively younger patients where better treatment outcomes are expected with lesser side effects. However, our study did not find that younger age is associated with favourable treatment response.

In the present study the odds of receiving treatment increased if the patient had MAC infection. As per study, the principal causative species for NTM-PD are members of MAC. Several groups in multiple countries have documented the increasing incidence of MAC related pulmonary disease [18]. Not all patients will require treatment initially but most of the patients will require treatment during the course of the disease. Interestingly, the first guidelines on the treatment of NTM lung disease in 2007 did not impact the decision to start treatment.

In the present study the patients who were initiated with treatment within 6 months or later did not differ in their time to culture conversion. One previous study [19] has noted that the waiting period between diagnosis and treatment of NTM-PD patients did not impact culture conversion in patients. There are reports of patients with infection by *M. abscessus*, MAC, and elderly patients having associations with treatment failure [20]. However, in the current study patients with MAC infection had higher odds of culture conversion. This could be due to predominance of MAC species.

An interesting finding in this study was the spontaneous culture conversion in 55% of patients who did not receive antibiotics with a relative lower relapse rate, as compared to culture conversion among 74% patients who received antibiotics with a relatively higher relapse rate. This highlights the importance of bronchial hygiene measures for improved clearance of mucus from the lungs [21]. These measures should be tried hopig for a spontaneous remission before initiation of antibiotics.

The current study has some limitations. It has a retrospective cohort design, and some missing clinical data, especially the susceptibility to drugs. The study has a limited sample size, which may skew the results and factors toward MAC, as it constitutes the majority of NTM infections. Additionally, a small sample size may not provide an accurate representation of the trends. Another limitation is the accuracy of data input, as we obtained our data by reviewing patients’ journals. However, since a digital patient file system was first used in the early 2000s, data entry during the initial years may not have been accurate, which could impact our findings. Therefore, we suggest that the results of this study need verification by large, multi-centre, prospective cohort studies.

## Conclusion

Our study found that the major species causing NTM pulmonary disease in Western Norway from 2000 to 2021 were MAC, *M. malmoense*, and *M. abscessus*. Increased age and presence of pulmonary comorbidity increased the risk of getting NTM pulmonary infection. Patients with high symptom scores, below the age of 65 and, MAC infection have higher odds of receiving treatment. A favourable treatment response was seen in 74% of patients given antibiotic treatment. Spontaneous remission was found in 55% of cases who did not receive treatment. Factors associated with favourable treatment response were not found. Further research is required to determine the extent to which these comorbidities may have influenced the presentation and diagnosis of NTM pulmonary infection and disease.

## Data Availability

All data produced in the present study are available upon reasonable request to the authors

